# Agreement between smartphone-based mobile sensing and actigraphy sleep metrics in young people with bipolar disorder

**DOI:** 10.64898/2026.02.20.26346722

**Authors:** Adrianna Lopaczynski, John Merranko, Jessica Mak, Mary Kay Gill, Tina R. Goldstein, Jennifer Fedor, Carissa Low, Jessica C. Levenson, Boris Birmaher, Danella M. Hafeman

**Affiliations:** University of Pittsburgh Medical Center; University of Pittsburgh School of Medicine, Department of Psychiatry; University of Pittsburgh School of Medicine, Department of Medicine

**Author notes:** Corresponding Author: Adrianna Lopaczynski. Disclosures:* The authors have no competing interests to disclose. This research was supported by following studies funded by the National Institute of Mental Health (NIMH): Predicting Recurrence of Mood in Patients with Bipolar Disorder (PROMPT-BD) RO1 MH126991 (PI: Birmaher).

**Keywords:** Bipolar disorder, adolescence, sleep, actigraphy, mobile sensing, digital phenotyping, sleep timing, mood recurrence

## Abstract

**Background:** Sleep disturbance is a core feature of bipolar disorder (BD) and often precedes mood recurrence, particularly in youth. Although actigraphy provides objective sleep measurement, it is limited by cost and monitoring duration. Passive smartphone-based mobile sensing offers a scalable alternative, but its validity in youths with BD is unclear.

**Methods:** Analyses included adolescents and young adults (ages 14–25) with BD-I/II from the PROMPT-BD study with at least four days of concurrent actigraphy and mobile sensing. Actigraphy-derived sleep metrics (total sleep time [TST], sleep onset, sleep offset, midsleep, wake after sleep onset [WASO]) were compared with smartphone-derived proxies (total offline time [TOT], onset, offset, midsleep, phone use after sleep onset [PASO]). Agreement was evaluated using root mean squared error (RMSE) and mixed-effects models. Zero-inflated negative binomial models examined associations between WASO and PASO. Sensitivity analyses tested robustness to missing data, smartphone use patterns, sleep window definitions, operating system, presence vs. absence of mood symptoms and anxiety, and weekend effects.

**Results:** Mobile sensing showed strong convergence with actigraphy for sleep timing and duration (standardized β = 0.54-0.75, all p < .0001). RMSEs were <21 minutes for onset, offset, midsleep, and TST, with strongest agreement for midsleep (RMSE = 14.8 minutes). Mobile sensing slightly overestimated sleep duration and estimated earlier timing. PASO underestimated WASO (RMSE = 48.8 minutes), but greater WASO significantly increased the odds of detecting any PASO (OR per 15 minutes = 1.35, p < .0001). Findings were robust across sensitivity analyses.

**Conclusions:** Passive smartphone-derived sleep metrics approximated actigraphy-based estimates of sleep timing and duration in youth with BD. Given the widespread availability of smartphones in this population, this supports their potential as scalable tools for monitoring circadian disruption and informing early intervention.

---

Bipolar Disorder (BD) is a serious mental illness, often onsetting during late adolescence, that is characterized by recurrent episodes of depression and (hypo)mania^1^. Not only are sleep disturbances diagnostic criteria of both the depressive and manic/hypomanic episodes that define BD, but often precede the development of mood disorders and mood recurrence in BD^2^. Insomnia and decreased need for sleep are among the most commonly experienced symptoms prior to a mood recurrence in youth with BD, suggesting their early identification in a clinical setting as essential to early intervention^3,4^. Furthermore, even during remission, individuals with BD have been found to have higher rates of sleep abnormalities compared to healthy controls^5,6^, indicating that circadian disruptions are a core feature of BD. Such sleep disturbances are detrimental to youth brain growth and cognitive development, academic performance, substance use, and exacerbate existing depression and suicidality^7–9^.

While much of the literature reporting BD-related sleep disturbances has been based on self-report measures, actigraphy has been shown to be a valid, objective instrument for estimating sleep and daily activity rhythms in the BD population, highly correlated with polysomnography^10^. Despite actigraphy being a relatively unobtrusive technique to measure several metrics of sleep based on motion, user confirmation (i.e. pressing a button on the watch), and ambient light, there are several challenges with implementing its use, including battery life, user compliance, and cost, which are critical to the monitoring of longitudinal changes in sleep associated with mood episodes^11,12^. In addition, research-grade actigraphy watches are generally worn for short periods of time, so they may not be the best tool for assessing mood-related changes over periods of months to years.

The use of smartphones to passively collect data regarding screen unlocks, activity, and location from embedded sensors (i.e., “mobile sensing”) has the potential to provide continuous, longitudinal, real-time indicators of sleep and activity that may improve symptom tracking and risk factor identification^13^. As compared to actigraphy, smartphones offer a more accessible and scalable sleep measurement system for youth, 95% of whom own a smartphone^14^. Most youth that have mobile devices check their phones before bed and upon waking, with nearly half of teens using their phones “almost constantly” and 93% using their phones at least “several times a day”^14^. Frequency of phone use may be even higher in the presence of sleep problems^15^. Previous studies have indicated that while mobile sensing methods overestimate sleep metrics in healthy adults^16^, they are highly feasible and acceptable in youth with mood disorders^17^. Mobile sensing metrics of sleep (e.g., late night phone use) have been found to correlate with actigraphy-based metrics in a large, community-based sample of adolescents^18^ and young adults^11^, and successfully capture clinical features of insomnia when compared to healthy controls^19^. However, to our knowledge, these relationships between mobile sensing and actigraphy-based measures of sleep have not yet been documented in young people with psychopathology, including BD, who have higher rates of sleep disruptions and irregular circadian rhythms.

Through the ongoing Predicting Recurrence of Mood in Patients with Bipolar Disorder (PROMPT-BD) study, our team has assessed mood, behavior, and sleep in adolescents and young adults with BD-I/II by using a combination of clinical assessments, actigraphy, and mobile sensing to measure factors associated with impending mood recurrences, such as sleep-activity rhythms and mobility. However, prior to assessing the degree to which sleep-related mobile sensing metrics predict mood symptoms, it is first critical to assess their validity relative to other objective, high-quality sleep assessment (e.g., through actigraphy). The primary goal of the current analysis is to examine, in this sample of youths and young adults with BD-I/II, how sleep-related mobile sensing metrics map onto relevant actigraphy measures; we also assess the degree to which these correlations are modified based on other factors, such as self-reported smartphone use patterns, missing mobile sensing data, and clinical factors.

## Methods & materials

### Participants

This study included 53 young people (14-25 years old, Table 2) with BD-I/II at the University of Pittsburgh, who were enrolled in PROMPT-BD prior to March 6, 2025, 23 of which were included for analyses (eTable 1). Participants were recruited from research registries (69%), outpatient clinics (26%), and referrals from other sources (5%), and were included if they were deemed currently in remission from BD-I/II (at least 8 weeks with minimal mood symptoms). Individuals with schizophrenia, autism, or IQ <70, as well as any condition that may interfere with data collection such as a serious medical or neurological illness, were excluded. Participants were assessed for eligibility after consenting but prior to their enrollment in study procedures, and later assessed at 6, 12, 18, and 24 months after their intake evaluation. The University of Pittsburgh’s Institutional Review Board approved all study procedures.

### Procedure

At intake, participants completed a diagnostic interview using either the Kiddie Schedule for Affective Disorders and Schizophrenia for School-Age Children–Present and Lifetime Version (KSADS-PL)^20^ (for participants <18 years old) or the Structured Clinical Interview (SCID)^21^ (for participants ≥18 years old). Psychosocial function was determined based on the Child Global Assessment Scale^22^ (<18) and Global Assessment of Functioning (≥ 18 years old). Psychiatric symptoms and exposure to treatment was assessed week-by-week using the Longitudinal Interval Follow-up Evaluation (LIFE), and quantified using the instrument’s Psychiatric Status Rating (PSR) scale^23^. The PSR uses numeric values based on the *DSM-IV* criteria and participant functioning. Scores ≤ 2 indicate euthymia, 3–4 for subthreshold symptoms, and ≥ 5 for full-threshold symptomatology. All assessments were administered by trained research staff who then presented to a psychiatrist or clinical psychologist to confirm diagnoses. The Hollingshead Scale^24^ was used to determine socioeconomic status.

Following the assessment, eligible participants were instructed to install the AWARE smartphone application on their personal Android or iOS devices. The AWARE platform was used to collect passive, real-time data on sleep and circadian rhythms over a 24-month period. Data were sampled multiple times per hour and included activity classification (e.g., sedentary), approximate device location, and screen unlocks.

In addition, participants were instructed to wear an actigraphy watch continuously for two weeks following the intake and each of the follow-up visits. During these periods, participants completed an electronic sleep diary twice daily to complement actigraphy data. At each visit, participants reported routine periods during the previous six months when they were away from their phone (e.g., school or sports practice) and whether they noticed the AWARE app or changed their behavior because of it.

### Instruments

*Actigraphy.* Sleep was assessed using both wrist-worn actigraphy and passive mobile sensing. Actigraphy-derived sleep metrics, which served as the reference standard for objective sleep measurement, included total sleep time (TST), sleep timing onset and offset timing, midsleep, sleep timing variability, wake after sleep onset (WASO), and number of awakenings. These measures were used to evaluate the validity of mobile sensing-derived sleep estimates.

*Mobile Sensing (AWARE).* Passive mobile sensing data were collected using the AWARE smartphone application to derive sleep metrics designed to match established actigraphy sleep constructs^25^. Passive sensing data were gathered using multiple smartphone sensors and calculated solely based on the recorded data points, meaning that no assumptions or inferred values are introduced, ensuring the derived features reflect only the available data. Features were extracted from these raw data using the Reproducible Analysis Pipeline for Data Streams (RAPIDS), enabling the calculation of behavioral measures across key domains. Mobile sensing data were aggregated into one-hour intervals, computing summary measures tracking movement, screen unlocks, and vehicle use. Mobile sensing proxies for sleep onset, midsleep, sleep offset, TST (Total Offline Time; TOT), and WASO (Phone After Sleep Onset; PASO) were calculated as described in Table 1. Primary analysis focused on a pre-defined sleep window (i.e., 9 p.m. to 11 a.m.) to decrease the potential for model error due to daytime hours away from the phone (e.g., at school or work).

**Table 1.** Actigraphy and Mobile Sensing Variable Definitions.

| **Actigraphy Variable (Reference)** | **Mobile Sensing Variable (Proxy)** | **Operational Definition** |
| --- | --- | --- |
| Sleep Onset | Screen-on timing, activity classification (sedentary, mobile, vehicle) | Sleep onset is defined as the first hour in the sleep window (9 PM – 11 AM) with no screen-on events, physical activity, or vehicle time. in this window. |
| Sleep Offset | Screen-on timing, activity classification (sedentary, mobile, vehicle) | Sleep offset is defined as the last hour within the sleep window (9 PM–11 AM) with no screen-on events, physical activity, or vehicle time. |
| Midsleep | Midsleep | Midsleep is defined as the midpoint between sleep onset and offset. |
| Wake After Sleep Onset (WASO) | Phone Use After Sleep Onset (PASO) | Sleep fragmentation was estimated using WASO, defined as the total duration of wakefulness occurring after sleep onset as measured by actigraphy, and PASO, defined as the total duration of phone screen–unlocked time occurring after sleep onset as measured via mobile sensing. |
| Total Sleep Time (TST) | Total Offline Time (TOT) | Sleep duration is estimated as the number of hours between “sleep onset” and “sleep offset”, after accounting for PASO. |

**Table 2.** Demographic and Clinical Characteristics.

|  |  |
| --- | --- |
| Intake Age, mean (standard deviation) | 21.3 (2.4) |
| Intake Diagnosis |  |
| Bipolar Disorder-I, n (%) | 18 (78.3%) |
| Bipolar Disorder-II, n (%) | 5 (21.7%) |
| Female, n (%) | 19 (82.6%) |
| Race |  |
| Black, n (%) | 1 (4.4%) |
| Asian, n (%) | 3 (13.0%) |
| White, n (%) | 14 (60.9%) |
| Multiracial, n (%) | 5 (21.7%) |
| Socioeconomic Status, mean (standard deviation) | 4.1 (0.9) |
| Global Assessment of Functioning, mean (standard deviation) | 71.7 (9.0) |
| Mood symptoms at time of assessment (PSR≥3 for depression and/or hypo/mania), n (%) | 11 (47.8)% |
| Comorbidity |  |
| Generalized Anxiety Disorder, n (%) | 12 (52.2%) |
| Attention Deficit Hyperactivity Disorder, n (%) | 5 (21.7%) |
| Psychosis, n (%) | 1 (4.4%) |
| Substance Use Disorder, n (%) | 8 (34.8%) |
| Family History of Bipolar Disorder, n (%) | 7 (30.4%) |

Because data were aggregated into one-hour intervals, sleep onset times were systematically positively biased, and sleep offset times were negatively biased. For example, if a participant last used their smartphone at 10:30 p.m., the first fully offline interval would be detected at 11:00 p.m., resulting in a 30-minute delay in estimated sleep onset. To address this bias, sleep onset times were corrected by subtracting 30 minutes and sleep offset times by adding 30 minutes, assuming a uniform distribution of true onset and offset times within each hour. Final TOT estimates reflect these bias corrections.

### Statistical Methods

Participants were included in this analysis if they completed intake and had at least one week with (1) at least four days of valid actigraphy data (i.e., 4 hours or more per day) and (2) concurrent mobile sensing data. Concurrent daily estimates of sleep onset, offset, midsleep, and TOT/TST were compared between mobile sensing and actigraphy measures via estimation of root mean squared error (RMSE), both computed across the full sample as well as aggregated within participant and averaged across the sample. Mixed linear models then regressed actigraphy measures on concurrent mobile sensing measures to estimate standardized slope coefficients, accounting for repeated measures within-participant via random intercepts. Because PASO exhibited substantial zero inflation and right skew, we used mixed-effects zero-inflated negative binomial regression to assess whether (1) WASO as measured by actigraphy was significantly associated with the odds that mobile sensing detected any PASO and (2) whether WASO minutes were significantly associated with the count of PASO minutes among observations with nonzero PASO. As sensitivity analyses, all analyses were repeated excluding days with incomplete mobile sensing data, excluding subjects based on reported smartphone usage traits (e.g., only evaluating PASO and WASO association among cases who reported using their phone when awakening after sleep onset), expanding the mobile sensing sleep detection interval by four hours (7 p.m. to 1 p.m.), and estimating RMSEs separately for iPhone and Android users. As an exploratory analysis, we interacted mixed model effects with a dichotomous weekend vs. weekday variable to assess whether associations between mobile sensing and actigraphy measures were moderated by weekend/weekday. We also similarly interacted effects with presence vs. absence of mood symptoms and generalized anxiety disorder (GAD) to assess whether associations between mobile sensing measures and actigraphy were moderated by symptomatology. Wilcoxon and Fisher’s exact tests compared the included actigraphy sample to the rest of the PROMPT-BD sample.

## Results

### Sample Characteristics

After excluding participants with insufficient actigraphy data (n=30), the sample consisted of 23 participants (5 Android users, 18 iOS users; mean age = 21.3 ± 2.4, 83% female, 61% Caucasian) with a median of 13 days (ranging 4-55 days) of follow-up with concurrently collected actigraphy and mobile sensing data. Other relevant sociodemographic and clinical characteristics for this sample are shown in Table 2. Comorbid disorders were common, with approximately half of the sample also meeting criteria for GAD (n=12, 52%). As compared to excluded participants (based on available actigraphy data), included participants were significantly more balanced on race (Fischer’s Exact Test p=.004); there were no significant differences on any other demographic or clinical characteristics (eTable 1). Only six participants recorded at least one day with incomplete mobile sensing data, and the total percentage of participant-days with incomplete data was only 2%; no participants were excluded due to insufficient mobile sensing data.

### Phone use questionnaire

As shown in eTable 2, most participants reported smartphone use very close to bedtime, with 70% indicating that they typically stopped using their phone less than five minutes before bed and the remaining 30% reporting cessation within 30 minutes of bedtime. Consistent with this pattern, smartphones were typically kept close during sleep periods, with 57% of participants reporting that they kept their phone in bed, and an additional 39% reporting that they kept it within reach of the bed. WASO smartphone engagement was also common, with 39% of the sample reporting that they checked their smartphone at least most of the time when awakening during the night. The entire sample reported that they first checked their smartphone within five minutes of awakening and/or used their smartphone as their alarm.

### Comparison of mobile sensing and actigraphy estimates of sleep timing and duration

As shown in Table 3, mobile sensing estimates of sleep onset, midsleep, and sleep offset were significantly earlier than corresponding actigraphy estimates, and mobile sensing tended to slightly overestimate total sleep time. However, overall agreement between modalities was high. Root mean square errors (RMSEs) for all four sleep metrics were under 21 minutes, with subject-aggregated RMSEs under 29 minutes. Agreement was strongest for midsleep, which showed the lowest error (RMSE = 14.8 minutes; subject-aggregated RMSE = 18.4 minutes). As shown in Figure 1, error in estimating sleep onset times was largest when mobile sensing indicated earlier sleep onset (e.g., 9pm; Figure 1a), whereas error in estimating sleep offset times was largest when mobile sensing indicated later sleep offset times (e.g., 11am; Figure 1c). In contrast, midsleep estimates showed consistently low error across the full range of midsleep values (Figure 1b). Mixed linear regressions estimated very strong associations between mobile sensing and actigraphy measures of sleep onset, midsleep, sleep offset, and sleep duration (TST/TOT), with standardized regression coefficients ranging from 0.54-0.75 (all p-values < 0.0001).

**Figure 1.**
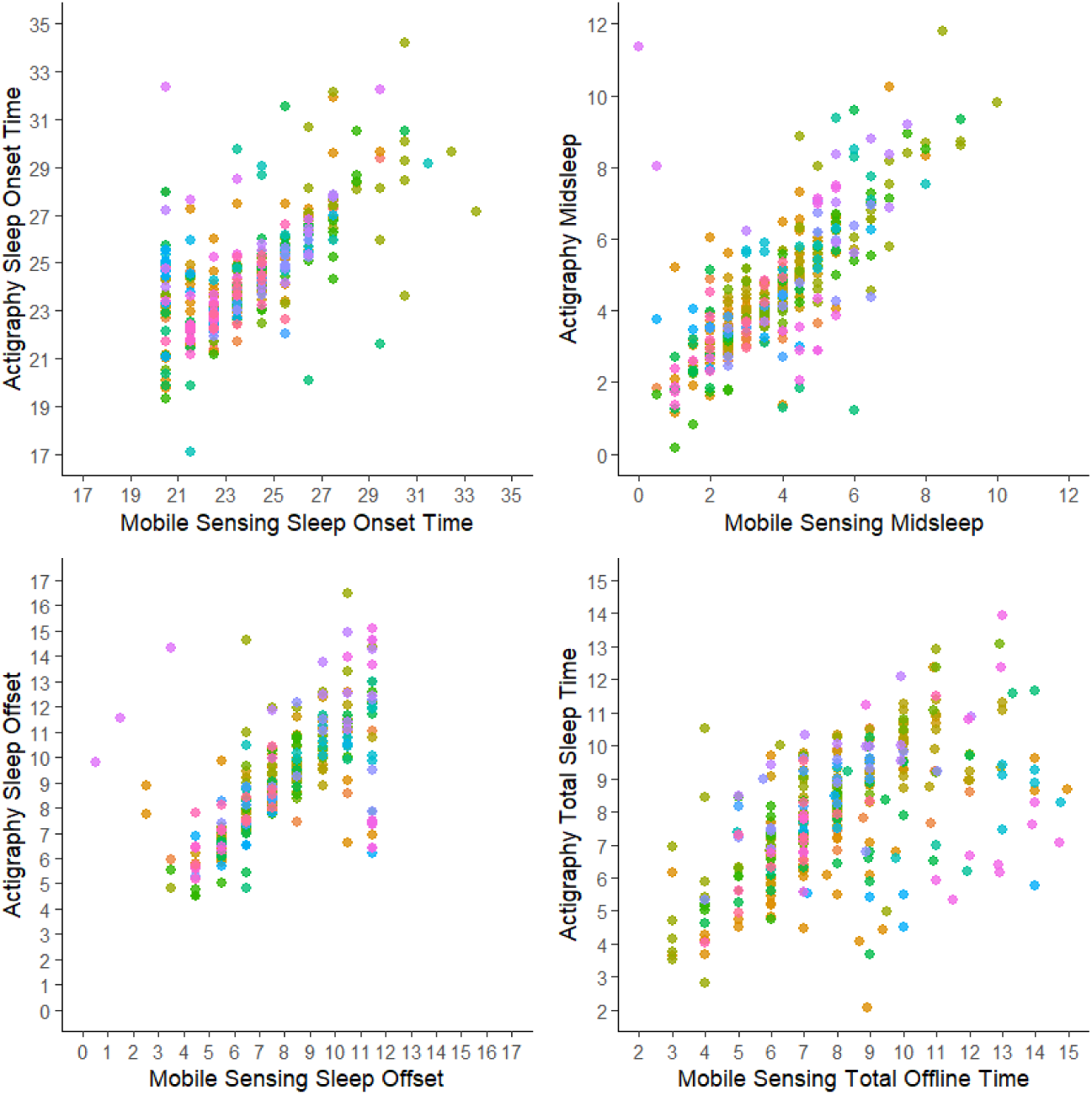
Mobile sensing metrics of sleep as compared to actigraphy. Points of the same color reflect repeated observations within-participant.

**Table 3.** Mobile sensing metrics of sleep as compared to actigraphy.

| Sleep Variable | AWARE Means | Actigraphy Means | RMSE | Subject-Aggregated RMSE | $\beta$ | 95% CI | | p-value |
| --- | --- | --- | --- | --- | --- | --- | --- | --- |
| Sleep Onset | 11:52 PM | 12:18 AM | 20.5 | 28.3 | 0.65 | 0.58 | 0.73 | <0.0001 |
| Midsleep | 3:48 AM | 4:36 AM | 14.8 | 18.4 | 0.75 | 0.68 | 0.82 | <0.0001 |
| Sleep Offset | 7:49 AM | 8:54 AM | 20.2 | 23.7 | 0.68 | 0.61 | 0.76 | <0.0001 |
| Total Sleep Time | 8.13 hours | 7.81 hours | 20.2 | 23.0 | 0.54 | 0.46 | 0.63 | <0.0001 |

RMSE = Root Mean Squared Error in minutes, $\beta$ = Linear Mixed Regression Coefficient (Standardized), CI = Confidence Interval

### Comparison of phone use after sleep onset and actigraphy wake after sleep onset

When regressing smartphone-derived PASO on actigraphy-derived WASO via mixed-effects zero-inflated negative binomial regression, actigraphy-derived WASO was strongly associated with the presence of any PASO (p-value < 0.0001; Figure 2a). Specifically, each 15-minute increase in WASO was associated with a 35% increase in the odds PASO detection (odds ratio = 1.35, 95% CI: 1.17-1.55), and each 30-minute increase in WASO was associated with an 82% increase in the odds of PASO detection (odds ratio = 1.82, 95% CI: 1.38-2.40). However, there was no significant relationship between minutes of WASO and PASO among observations with nonzero PASO (incident rate ratio per 15 minutes of WASO = 1.13, 95% CI: 0.90-1.43, p = 0.29; Figure 2b). Not surprisingly, smartphone derived estimates of PASO substantially underestimated actigraphy-derived WASO, with RMSE = 48.8 minutes and subject-aggregated RMSE = 46.7 minutes.

**Figure 2.**
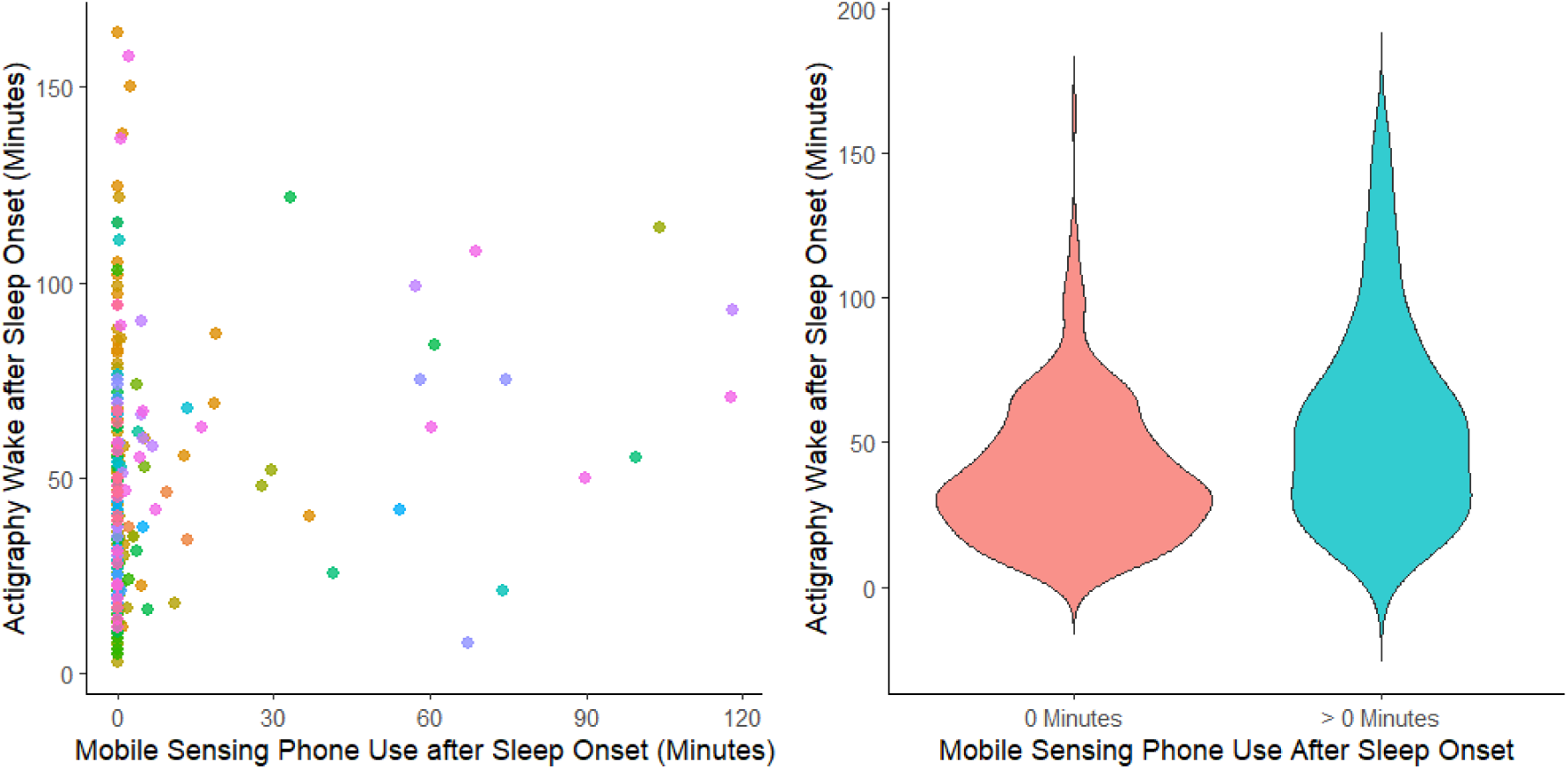
Association between Actigraphy Wake after Sleep Onset and Mobile Sensing Phone Use after Sleep Onset. Points of the same color reflect repeated observations within-participant.

### Sensitivity Analyses

*Effect of Incomplete Mobile Sensing Data.* As shown in Figure 3, excluding days with incomplete mobile sensing data eliminated several outlying points with large errors. However, as shown in eTable 3, RMSE and mixed linear regression results did not appreciably change.

**Figure 3.**
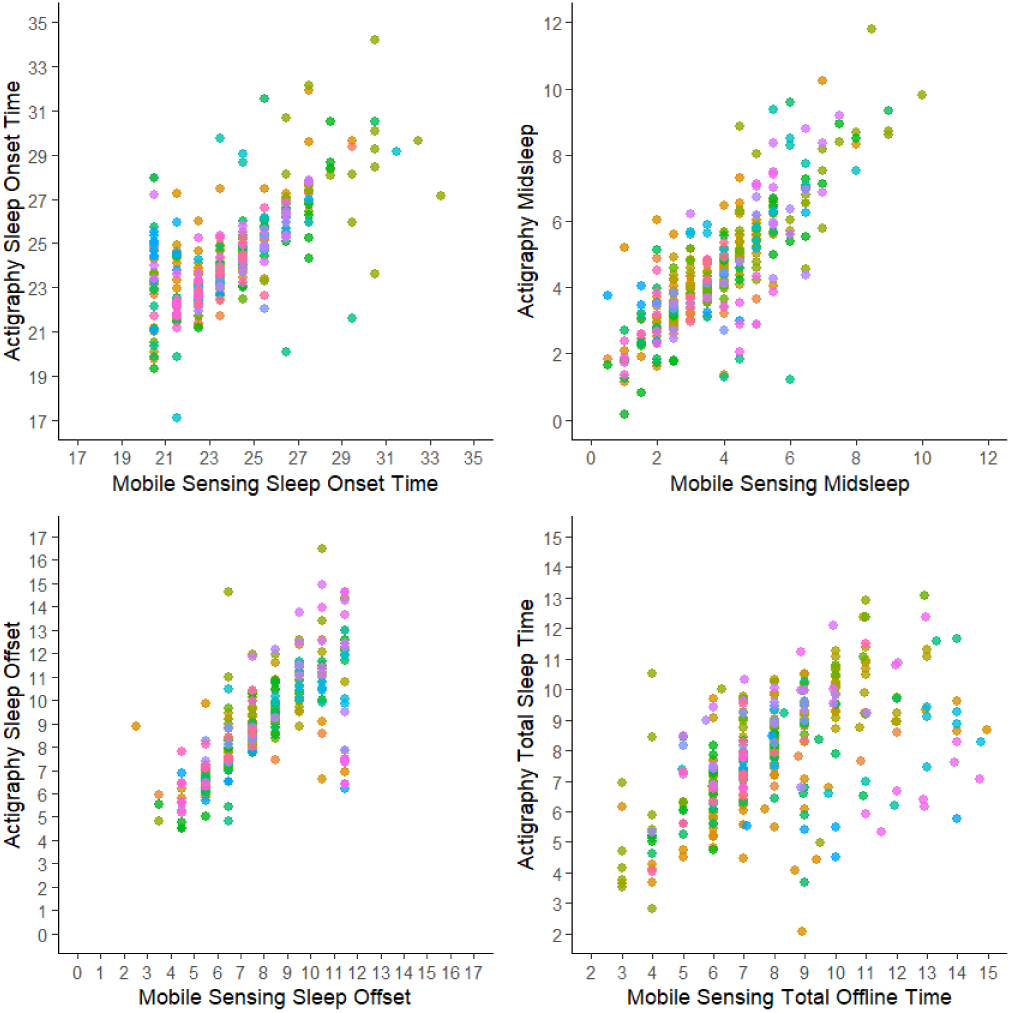
Complete Mobile Sensing Days Only. Points of the same color reflect repeated observations within-participant.

*Effect of Differences in Self-Reported Smartphone Usage.* Because all subjects reported that they checked their smartphones within five minutes of awakening or used them as an alarm, we were unable to perform a sensitivity analysis to determine whether mobile sensing estimates of sleep offset were affected by this factor. However, we were able to perform sensitivity analyses regarding the estimation of sleep onset, midsleep, and sleep duration among 16 participants who reported they stopped using their smartphones less than five minutes before bedtime. As shown in eTable 4, results did not appreciably change. When reevaluating the association between PASO and WASO among nine participants who reported checking their phone when awakening in the middle of the night either “most of the time” or “pretty much every time “, results indicated a slightly stronger association between WASO and presence of any PASO. Specifically, each 15-minute increase in WASO in this subsample was associated with a 37% increase in the odds PASO detection (odds ratio = 1.37, 95% CI: 1.11-1.68), and each 30-minute increase in WASO was associated with an 87% increase in the odds of PASO detection (odds ratio = 1.87, 95% CI: 1.24-2.82).

*Effect of Expanding Time Window Constraints.* We assessed the impact of broadening our pre-defined sleep window (from 9p.m.-11a.m. to 7 p.m.-1 p.m.) to allow for a greater range of variability in sleep problems. However, this increased overall sleep feature prediction error and, importantly, led to more outliers (eFigure 1, eTable 5).

*Sensitivity Analyses by Smartphone Operating System*. RMSEs and subject-aggregated RMSEs were smaller among iOS users than Android users for all sleep features estimated (eTable 6). While this may suggest that mobile sensing features estimated on iOS devices estimate sleep features with less error than Android devices, it is important to consider that only five participants (22%) used Android devices in our sample.

### Exploratory analysis of weekend/weekday moderation effects

Associations between mobile sensing and actigraphy measures of sleep onset, midsleep, offset, and sleep duration (TST/TOT) were all significantly moderated by weekend vs. weekday (interaction effect p-values < 0.04). As shown in eTable 7, associations were significantly stronger when the next day was a Saturday or Sunday, with estimated standardized regression coefficient differences between 0.13-0.20 (small-to-medium size interaction effects) for nights leading into weekend vs. weekdays. However, the association between actigraphy WASO and mobile sensing PASO was not significantly moderated by this effect (interaction p-value = 0.24).

### Exploratory analysis of mood symptom and generalized anxiety disorder effects

Only nine participants had at least one week with subthreshold or worse depression concurrent to mobile sensing and actigraphy data (median duration = 1 week), and only four participants had at least one week with subthreshold or worse hypomania concurrent to mobile sensing and actigraphy data (median duration = 1 week), so moderation models used the presence of *any* mood symptoms (n=11) as an interaction variable. Results indicated that the associations between mobile sensing and actigraphy estimates of midsleep and sleep offset were significantly stronger if participants experienced concurrent mood symptoms (interaction p-values = 0.003 and 0.005, respectively; eTable 8), with estimated standardized regression coefficient differences of 0.21 and 0.24, respectively (small-to-medium sized interaction effects). However, even in the absence of mood symptoms, we observed strong associations between these mobile sensing measures and relevant actigraphy metrics (p<0.0001).

GAD was the only comorbid disorder with sufficient representation in the sample (n>10) to similarly assess for interaction effects. Results indicated that the associations between mobile sensing and actigraphy estimates of sleep onset and midsleep were present regardless of GAD (p<0.0001), but were found to be significantly stronger among participants with GAD (interaction p-values = 0.03 and 0.02, respectively), with estimated standardized regression coefficient differences of 0.17 and 0.19, respectively (small-to-medium sized interaction effects; eTable 8). The association between actigraphy WASO and mobile sensing PASO was not significantly moderated by the presence of any concurrent mood symptoms (p-value = 0.07) or generalized anxiety disorder (p-value = 0.22).

## Discussion

The present study provides initial evidence that mobile sensing-derived sleep metrics can approximate key actigraphy-based sleep measures in youths and young adults with BD, supporting their potential utility as scalable, real-time indicators of sleep disturbance and changes in sleep timing. Across the sample, mobile sensing-based sleep variables showed convergence with actigraphy-derived estimates, suggesting that passive smartphone data may capture meaningful aspects of sleep-wake behavior in this population. While mobile sensing metrics do not map perfectly onto all actigraphy measures, their ability to approximate core sleep parameters is notable, particularly given their low participant burden and continuous availability. Overall, responses to the phone use questionnaire suggest that smartphones were highly integrated into sleep timing/behaviors and therefore have strong potential as a sleep tracking tool in this sample of young people with BD.

Importantly, these findings align with prior work demonstrating that behavioral proxies derived from personal devices can reflect clinically relevant sleep patterns and circadian disruptions associated with affective instability^26^. Sleep disruption is among the most robust early warning signs of both manic and depressive episodes, and the ability to passively detect such changes in naturalistic settings could substantially enhance early detection of mood episodes. Given the impact of repeated mood episodes on outcomes such as psychosocial functioning, substance abuse, and suicidal thoughts and behaviors^7–9^, particularly if these episodes are not addressed in a timely manner, such an early warning signal could have important implications for treatment of BD. In addition, sleep changes can be an important target of early intervention, as there is a strong evidence-base for therapeutic interventions (e.g., interpersonal social rhythms therapy^27–29^, CBT for insomnia^30–32^) that can improve sleep. Consistent with prior work^16,19^, our findings suggest that mobile sensing is particularly well-suited for capturing sleep disturbances among symptomatic BD youth. Mobile-sensing derived sleep metrics were significantly associated with actigraphy values regardless of mood state, but even more pronounced among participants with active mood symptoms and comorbid GAD, underscoring sensitivity to clinically meaningful sleep disruption. Thus, an important future direction will be to develop just-in-time interventions for sleep in this high-risk population, based on data obtained from mobile sensing.

When identifying sleep periods using mobile sensing data, we limited our “sleep window” of 9:00 PM to 11:00 AM. This use of predefined cutoffs in mobile sensing sleep variables has the limitation of excluding potentially informative variability. However, allowing “sleep time” throughout the day may lead to misidentification of time away from the smartphone (e.g., for work or school) as sleep. Indeed, when we conducted a sensitivity analysis that broadened the proposed sleep window, model performance declined and the number of outliers increased. In this way, the cutoffs ultimately reduce noise inherent in passive data streams and increase clinical interpretability.

When examining weekday versus weekend moderation in phone usage, contrary to expectations that more structured and consistent weekday behavior would yield stronger associations between mobile sensing and actigraphy measures, results indicated stronger correspondence on weekends (although effects for both were large). One possible explanation is that weekends provide richer and more continuous phone usage data, reflecting fewer externally imposed constraints (e.g., work or academic schedules) and greater engagement with personal devices. Alternatively, less disciplined or more variable phone use patterns on weekends may paradoxically enhance the sensitivity of mobile sensing algorithms to detect sleep-wake transitions, particularly during a “leisure window” in which behavior more closely reflects intrinsic circadian tendencies. These findings highlight the importance of contextualizing mobile sensing data within patterns of daily life and suggest that variability itself may be informative rather than a source of noise.

Several limitations should be considered when interpreting these findings. First, the analyzed sample was small and relatively homogeneous, limiting generalizability across broader and more diverse populations. Especially when considering the impact of other factors on sleep, such as comorbid disorders, environmental factors, medications, and substance use, this dataset did not have the statistical power to test more than the modifying effects of current mood symptoms and comorbid GAD in this population of BD youth. Second, while mobile sensing metrics approximated several actigraphy-based sleep measures, they did not accurately capture all dimensions of sleep quality, most notably WASO. WASO is closely related to the construct of sleep efficiency, a well-established marker of sleep quality and has been consistently associated with depressive symptomatology^33^. Finally, the median duration of overlapping mobile sensing and actigraphy data was approximately two weeks, limiting the ability to examine seasonal effects, longer-term trajectories, or within-person longitudinal dynamics.

Future research could expand upon this work by extending follow-up periods to enable examination of whether within-person deviations in mobile sensing-estimated sleep timing, variability, or regularity prospectively predict manic, hypomanic, or depressive episodes, and whether these signals precede subjective symptom change. Including machine learning approaches that incorporate individualized baselines and adaptive sleep windows may enhance sensitivity while preserving interpretability. Future studies should also test the clinical utility of embedding these metrics within just-in-time adaptive intervention frameworks, evaluating whether algorithm-triggered sleep-focused strategies (e.g., prompts to regularize bed/wake times, reduce nighttime screen exposure, or initiate brief CBT-I informed exercises) can mitigate emerging circadian disruption and reduce mood episode recurrence. Finally, ethical and implementation considerations, including data privacy, algorithmic transparency, and equitable access, are essential to address as mobile-sensing transitions from research settings to clinical care. Collectively, these research advancements will help to clarify whether passive sleep monitoring can become a scalable tool for precision prevention.

Overall, these analyses provide early support for the feasibility and clinical relevance of mobile-sensing based sleep metrics in adolescents and young adults with BD. Mobile-sensing holds significant promise as an accessible, real-time tool for monitoring sleep that may identify targets for intervention and predict mood recurrence. With further refinement, validation, and longitudinal investigation, these approaches may play a critical role in advancing early detection and preventative care in young people with BD.

## Data Availability

All data produced in the present study are available upon reasonable request to the authors.

## eSupplement

**eTable 1.**
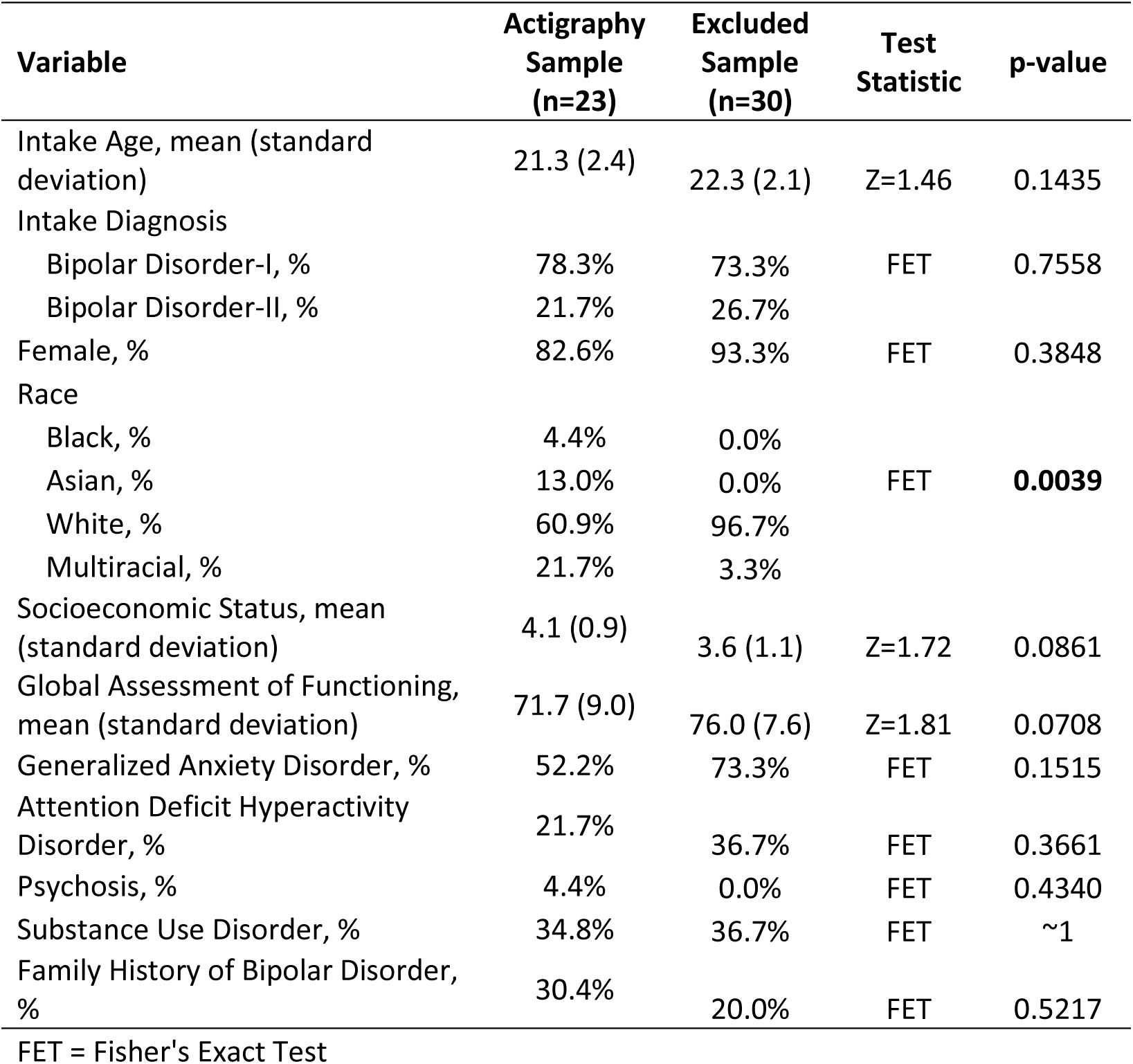
Comparing Included vs. Excluded Sample on Demographic and Clinical Characteristics.

**eTable 2.**
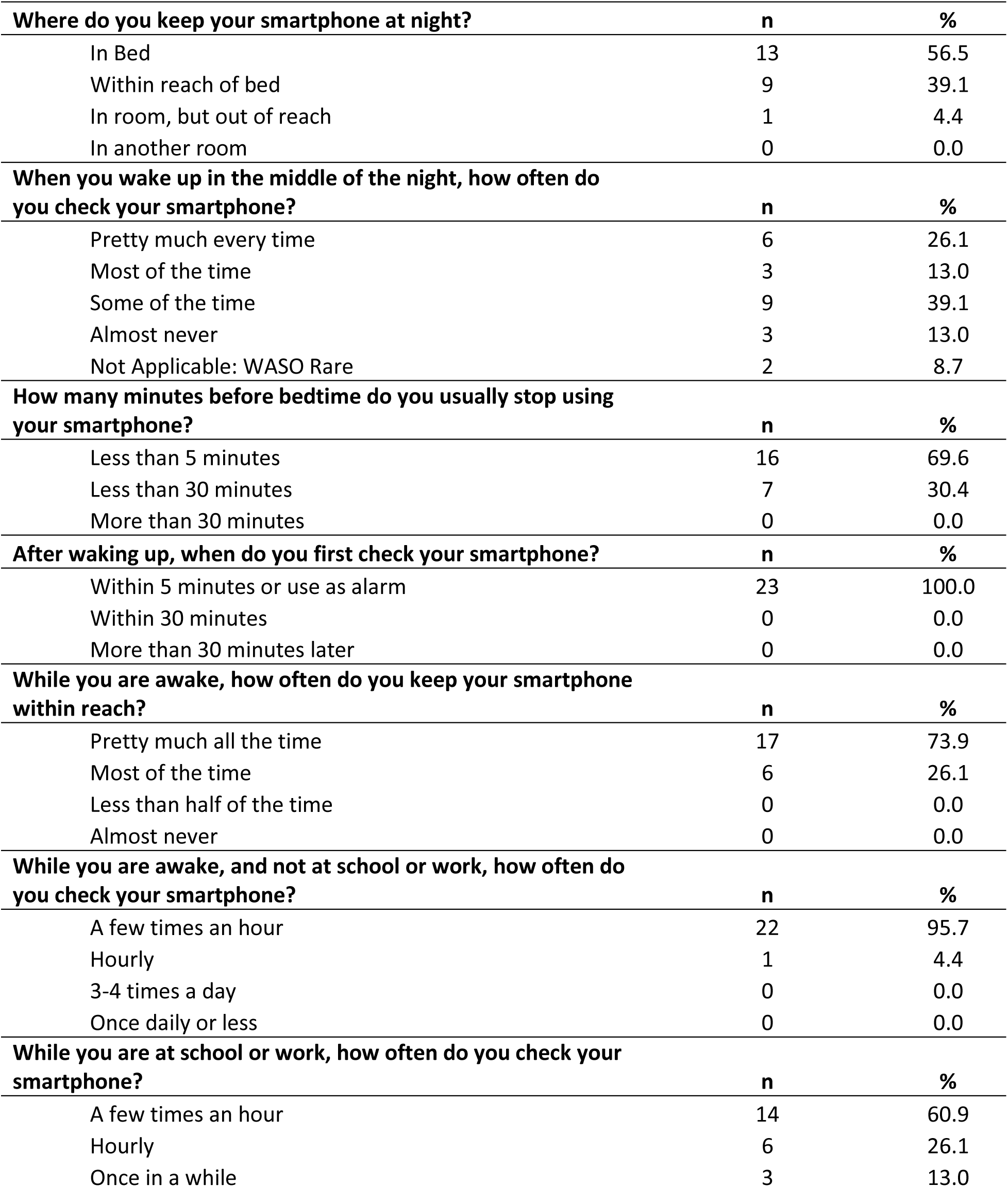
Phone Use Questionnaire.

| **Where do you keep your smartphone at night?** | **n** | **%** |
| --- | --- | --- |
| In Bed | 13 | 56.5 |
| Within reach of bed | 9 | 39.1 |
| In room, but out of reach | 1 | 4.4 |
| In another room | 0 | 0.0 |
| **When you wake up in the middle of the night, how often do you check your smartphone?** | **n** | **%** |
| Pretty much every time | 6 | 26.1 |
| Most of the time | 3 | 13.0 |
| Some of the time | 9 | 39.1 |
| Almost never | 3 | 13.0 |
| Not Applicable: WASO Rare | 2 | 8.7 |
| **How many minutes before bedtime do you usually stop using your smartphone?** | **n** | **%** |
| Less than 5 minutes | 16 | 69.6 |
| Less than 30 minutes | 7 | 30.4 |
| More than 30 minutes | 0 | 0.0 |
| **After waking up, when do you first check your smartphone?** | **n** | **%** |
| Within 5 minutes or use as alarm | 23 | 100.0 |
| Within 30 minutes | 0 | 0.0 |
| More than 30 minutes later | 0 | 0.0 |
| **While you are awake, how often do you keep your smartphone within reach?** | **n** | **%** |
| Pretty much all the time | 17 | 73.9 |
| Most of the time | 6 | 26.1 |
| Less than half of the time | 0 | 0.0 |
| Almost never | 0 | 0.0 |
| **While you are awake, and not at school or work, how often do you check your smartphone?** | **n** | **%** |
| A few times an hour | 22 | 95.7 |
| Hourly | 1 | 4.4 |
| 3-4 times a day | 0 | 0.0 |
| Once daily or less | 0 | 0.0 |
| **While you are at school or work, how often do you check your smartphone?** | **n** | **%** |
| A few times an hour | 14 | 60.9 |
| Hourly | 6 | 26.1 |
| Once in a while | 3 | 13.0 |

**eTable 3.** Sensitivity Analysis: Complete Mobile Sensing Days Only.

| Sleep Variable | AWARE Means | Actigraphy Means | RMSE | Subject-Aggregated RMSE | $\beta$ | 95% CI | | p-value |
| --- | --- | --- | --- | --- | --- | --- | --- | --- |
| Sleep Onset | 11:51 PM | 12:18 AM | 19.4 | 30.6 | 0.66 | 0.59 | 0.73 | <0.0001 |
| Midsleep | 3:48 AM | 4:36 AM | 12.9 | 18.7 | 0.76 | 0.69 | 0.82 | <0.0001 |
| Sleep Offset | 7:51 AM | 8:54 AM | 18.4 | 24.3 | 0.69 | 0.62 | 0.76 | <0.0001 |
| Total Sleep Time | 8.11 hours | 7.81 hours | 20.8 | 26.5 | 0.52 | 0.44 | 0.60 | <0.0001 |

RMSE = Root Mean Squared Error in minutes, $\beta$ = Linear Mixed Regression Coefficient (Standardized), CI = Confidence Interval

**eTable 4.** Sensitivity Analysis: participants who reported they stopped using their smartphones less than five minutes before bedtime (n=16)

| Sleep Variable | RMSE | Subject-Aggregated RMSE | $\beta$ | 95% CI | | p-value |
| --- | --- | --- | --- | --- | --- | --- |
| Sleep Onset | 25.5 | 31.2 | 0.65 | 0.56 | 0.74 | <0.0001 |
| Midsleep | 16.4 | 18.1 | 0.74 | 0.66 | 0.81 | <0.0001 |
| Total Sleep Time | 24.4 | 24.1 | 0.57 | 0.47 | 0.66 | <0.0001 |

RMSE = Root Mean Squared Error in minutes, $\beta$ = Linear Mixed Regression Coefficient (Standardized), CI = Confidence Interval

**eTable 5.** Expanding mobile sensing sleep detection interval to 7 p.m. – 1 p.m.

| Sleep Variable | RMSE | Subject-Aggregated RMSE | $\beta$ | 95% CI | | p-value |
| --- | --- | --- | --- | --- | --- | --- |
| Sleep Onset | 61.5 | 78.6 | 0.16 | 0.08 | 0.23 | <0.0001 |
| Midsleep | 24.7 | 30.0 | 0.35 | 0.25 | 0.45 | <0.0001 |
| Sleep Offset | 23.9 | 28.5 | 0.44 | 0.34 | 0.53 | <0.0001 |
| Total Sleep Time | 65.9 | 76.4 | 0.16 | 0.06 | 0.26 | 0.0024 |

RMSE = Root Mean Squared Error in minutes, $\beta$ = Linear Mixed Regression Coefficient (Standardized), CI = Confidence Interval

**eTable 6.** Sensitivity analyses by smartphone operating system.

| Sleep Variable | Operating System | RMSE | Subject-Aggregated RMSE |
| --- | --- | --- | --- |
| Sleep Onset | Android (n=5) | 42.5 | 58.5 |
| Midsleep |  | 22.5 | 30.2 |
| Sleep Offset |  | 20.5 | 29.2 |
| Total Sleep Time |  | 38.3 | 39.7 |
| Wake after Sleep Onset |  | 51.9 | 54.8 |
| Sleep Onset | iOS (n=18) | 18.1 | 19.9 |
| Midsleep |  | 14.2 | 15.1 |
| Sleep Offset |  | 20.2 | 22.2 |
| Total Sleep Time |  | 18.3 | 18.4 |
| Wake after Sleep Onset |  | 48.6 | 44.4 |

RMSE = Root Mean Squared Error in minutes

**eTable 7.** Exploratory analysis of weekend/weekday moderation effects.

| Sleep Variable | Weekend/Weekday | $\beta$ | 95% CI | | p-value | Type 3 Interaction p-value |
| --- | --- | --- | --- | --- | --- | --- |
| Sleep Onset | Weekend | 0.79 | 0.66 | 0.91 | <0.0001 | 0.0229 |
|  | Weekday | 0.62 | 0.53 | 0.72 | <0.0001 |  |
| Midsleep | Weekend | 0.83 | 0.72 | 0.94 | <0.0001 | 0.0399 |
|  | Weekday | 0.70 | 0.62 | 0.78 | <0.0001 |  |
| Sleep Offset | Weekend | 0.79 | 0.66 | 0.92 | <0.0001 | 0.0324 |
|  | Weekday | 0.63 | 0.54 | 0.71 | <0.0001 |  |
| Total Sleep Time | Weekend | 0.67 | 0.53 | 0.81 | <0.0001 | 0.0162 |
|  | Weekday | 0.48 | 0.38 | 0.57 | <0.0001 |  |

$\beta$ = Linear Mixed Regression Coefficient (Standardized), CI = Confidence Interval

**eTable 8.** Exploratory analysis of weekend/weekday moderation effects.

| **Sleep Variable** | **Moderator** | **$\beta$** | **95% CI** |  | **p-value** | **Type 3 Interaction p-value** |
| --- | --- | --- | --- | --- | --- | --- |
| Sleep Onset | Concurrent Mood Symptoms* | 0.76 | 0.63 | 0.89 | <0.0001 | 0.2215 |
|  | No Mood Symptoms | 0.67 | 0.58 | 0.75 | <0.0001 |  |
| Midsleep | Concurrent Mood Symptoms* | 0.89 | 0.78 | 1.00 | <0.0001 | 0.0026 |
|  | No Mood Symptoms | 0.68 | 0.59 | 0.76 | <0.0001 |  |
| Sleep Offset | Concurrent Mood Symptoms* | 0.83 | 0.70 | 0.97 | <0.0001 | 0.0052 |
|  | No Mood Symptoms | 0.59 | 0.49 | 0.69 | <0.0001 |  |
| Total Sleep Time | Concurrent Mood Symptoms* | 0.54 | 0.39 | 0.70 | <0.0001 | 0.9535 |
|  | No Mood Symptoms | 0.54 | 0.43 | 0.64 | <0.0001 |  |
| Sleep Onset | GAD | 0.71 | 0.62 | 0.80 | <0.0001 | 0.0330 |
|  | No GAD | 0.54 | 0.41 | 0.67 | <0.0001 |  |
| Midsleep | GAD | 0.80 | 0.72 | 0.88 | <0.0001 | 0.0205 |
|  | No GAD | 0.60 | 0.46 | 0.75 | <0.0001 |  |
| Sleep Offset | GAD | 0.68 | 0.59 | 0.77 | <0.0001 | 0.8396 |
|  | No GAD | 0.70 | 0.56 | 0.84 | <0.0001 |  |
| Total Sleep Time | GAD | 0.52 | 0.41 | 0.62 | <0.0001 | 0.3592 |
|  | No GAD | 0.60 | 0.45 | 0.75 | <0.0001 |  |

$\beta$ = Linear Mixed Regression Coefficient (Standardized), CI = Confidence Interval

**eFigure 1.**
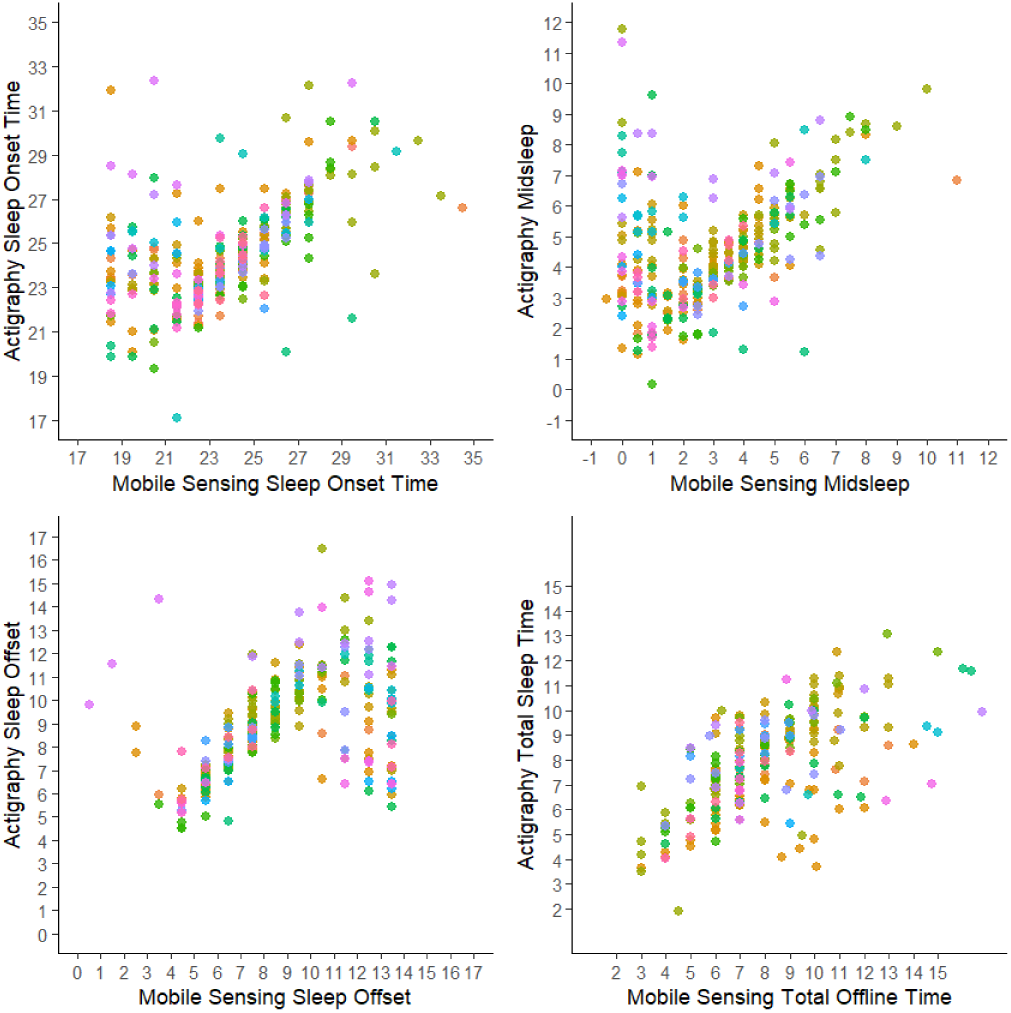
Expanding mobile sensing sleep detection interval to 7 p.m. – 1 p.m.

